# Development and Clinical Validation of a Miniaturized Finger Probe for Bedside Hemodynamic Monitoring

**DOI:** 10.1101/2022.08.26.22279246

**Authors:** Tuukka Panula, Jukka-Pekka Sirkiä, Tero Koivisto, Mikko Pänkäälä, Teemu Niiranen, Ilkka Kantola, Matti Kaisti

## Abstract

**Background:** Hypertension, or elevated blood pressure (BP), is a marker for many cardiovascular diseases and can lead to life-threatening conditions such as heart failure, coronary artery disease, and stroke. BP monitoring using a traditional arm cuff device is often inconvenient and possibly painful in long-term use, i.e. during sleep.

**Methods:** We present a miniature cuffless tonometric finger probe system, that uses the oscillometric method to measure blood pressure (BP). Our approach uses a motorized press that is used to apply pressure to the finger tip to measure BP.

**Results:** We verified the functionality of the device in a clinical trial (n=43) resulting in systolic (SBP) and diastolic (DBP) pressures ((*mean* ± *SD*) mmHg) of (−3.5 ± 8.4) mmHg and (−4.0 ± 4.4) mmHg, respectively. Comparison was made with manual auscultation (n=26) and automated cuff oscillometry (n=18). In addition to BP, we demonstrated the ability of the device to assess arterial stiffness (n=18) and detect atrial fibrillation (n=6).

**Conclusions:** We were able to introduce a sufficiently small device that could be used for convenient ambulatory measurements and worn during sleep with minimal discomfort. Lastly, we evaluate methods to further develop the concept and discuss future directions.

**Plain language summary:** Traditional cuff-based automated BP instruments are mainly suitable for individual measurements and are often inconvenient to use. Patients in a hospital ward are typically monitored with many systems and devices. Our aim is to develop a blood pressure measurement technology that could be integrated into a finger-worn pulse oximeter. This way the need for a bulky and inconvenient arm cuff could be eliminated and the number of needed monitoring devices could be reduced. The proposed technology is especially suitable for pseudo-continuous BP measurement, that is, taking automated measurements periodically, enabling BP trend tracking and analysis. In this study, we clinically validate our technology for blood pressure monitoring and further demonstrate its potential to measure arterial stiffness and detect atrial fibrillation.

## 1 Introduction

Untreated hypertension is the greatest single contributor for cardiovascular disease and sudden cardiac death. Uncontrolled high blood pressure (BP) can lead to disability, poor quality of life, and fatal heart attack or stroke [31]. With changing global lifestyle, hypertension is affecting ever-younger individuals and its prevalence is increasing. [8]. Currently, the trend in BP monitoring devices is going towards wearable technologies and ease-of-use [25]. Current hypertension guidelines also recommend that blood pressure is measured regularly and frequently (instead of random spot measurement)[17], despite traditional BP devices with an inflatable arm cuff being uncomfortable, especially in long-term use.

Most wearable BP measurement devices use pulse wave propagation or pulse wave analysis methods to measure BP [18, 19, 29]. These devices often require initial arm cuff calibration using a brachial cuff device. Moreover, these methods typically lack clinical validation and special situations and populations are seldom considered. Our device relies on the oscillometric method to acquire spot BP measurements and it does not require any calibration [9]. Similar techniques have been introduced before, but they require a bulky external pneumatic unit for pressure generation, or are based on user actuation [20, 26, 6].

In our previous study we presented a tabletop tonometric instrument for measuring blood pressure from the fingertip[23]. Operation of the device was validated (n=33) against an automated arm cuff device (Omron M3) resulting in systolic (SBP) and diastolic (DBP) pressures ((*mean* ± *SD*) mmHg) of (−0.9 ± 7.3) mmHg and (−3.3 ± 6.6) mmHg, respectively. We have also proposed a miniaturized version of the device in another study[24].

In this study, our main objective was to validate the proposed device according to the international BP standards. This involves comparing it to manual auscultation, the gold standard method for non-invasive monitoring of BP[3]. The standards used for most BP devices are maintained by the US Association for the Advancement of Medical Instrumentation (AAMI), the European Society of Hypertension Working Group on Blood Pressure Monitoring (ESH), British Hypertension Society (BHS) and the International Organization for Standardization (ISO) [30, 4, 21].

Our secondary objective was to miniaturize the technology. The main motivation for miniaturization is to be able to integrate the technology to a pulse oximeter, eliminating the need for an inconvenient brachial cuff and acquiring both the blood pressure and blood oxygenation from a sinle finger probe. In addition, we assessed the device’s capability to measure arterial stiffness and atrial fibrillation.

Thirdly, we wanted to demonstrate the device’s capability to recognize cardiac arrhythmias. Atrial fibrillation (AF) is known to affect the accuracy of the oscillometric method[22]. In the case of AF, the BP results should be examined critically. Inaccurate BP values can lead to incorrect treatment or even lack of it. Consequently, built-in AF detection algorithms are a requirement of a well performing BP monitor [28].

## 2 Methods

### 2.1 Device development

#### 2.1.1 Electronics

The device uses the nRF52840 (Nordic Semiconductor, Norway) microcontroller unit (MCU) dongle. The MCU was selected because of its high performance ARM Cortex M4 core and built-in USB capabilities. A barometric pressure sensor (BMP180, Bosch Sensortec, Germany) is connected to the MCU via I^2^C connection. A DC motor is used to apply pressure to the finger. The MCU is interfaced to the motor via DRV8837 motor driver (Texas Instruments, USA) H-bridge that enables bidirectional motor control. The MCU also monitors the two limit switches (SSCQ110100, Alps Alpine, Japan) located on a separate PCB. The MCU is connected to a laptop computer via USB.

#### 2.1.2 Mechanics

The mechanics of the instrument house the electronic components, provide a mechanism for applying controlled pressure to the finger as well as a structural base for the tonometric pressure sensor. The enclosure was 3D printed using a selective laser sintering process. A standard pressure sensor is modified by prying off its protective metal lid, and placing an air-filled cushion on top of it. A cylindrical piston is placed onto the cushion and it acts as the tonometer probe. The piston consists of two pieces attached together from the ends of a threaded shaft, enabling piston extension and distension in case the protrusion height of the piston has to be adjusted. A DC motor (generic N20 type) followed by a reduction gear train is placed in the inner supporting enclosure. A worm gear assembly further reduces the rotational speed and changes the angle of rotation. The whole transmission is encased in an inner support structure. A type of folding press system is used to apply pressure to the finger. The portion making contact to the finger is made from a flexible 3D printing filament. In order to make sure the pressing device does not move out of its range, the hinge is equipped with limit switches for the lower and upper boundaries. This also prevents possible injuries that could be caused by excessive pressure to the finger.

#### 2.1.3 Software

The device firmware is written in the C programming language using the nRF5 software development kit (SDK). The main functionalities of the firmware are sampling the pressure sensor with sampling frequency of 128 Hz, and controlling the pressure-generating motor.

The measurement is controlled with a graphical user interface (GUI) written in Python. The application connects to the device through a USB serial connection, which is used to send commands and receive data. The application plots the incoming data in real-time. The user interface allows the user to change the settings of the device, such as the speed of the motor and the maximum allowed pressure, as well as start and stop the measurement. Once a measurement has been completed, the received data together with the used device settings are stored in a comma-separated values (CSV) file.

### 2.2 Data analysis

Data analysis was performed on MATLAB R2019a (Mathworks, USA). Each measurement was stored in a separate CSV file and processed individually.

Blood pressure was computed by first subtracting the atmospheric pressure from the signal to set the base level for the measurement. The data are filtered with a pass-band of 1 to 10 Hz and the filtered signal is Hilbert transformed. A peak detection algorithm followed by 8th order polynomial fitting is applied. The resulting bell-shaped curve is called an oscillometric waveform envelope (OMWE). Mean arterial pressure (MAP) is found at the point on the pressure curve that corresponds with the maximum of OMWE. This also represents zero transmural pressure, *P*_*t*_ = 0. Transmural pressure is defined as the difference between intra-arterial and external applied pressure. DBP is found at the pressure level corresponding to 80% of maximum oscillations at the left side of OMWE. [23]

Systolic pressure is calculated from DBP and MAP using the following equation:

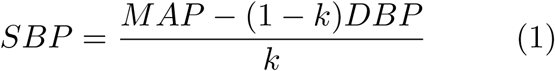

We used two methods to calculate the parameter *k*, resulting in two different values SBP. In the standard method, k was defined as *k* = 0.4 for all subjects. For a patient-specific approach, we used ASI as an additional parameter in *k*: *k* = 0.4 + *l*, where 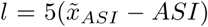. 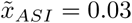 is the median of the calculated ASIs in the subject group with measured PWV (n = 18).

Arterial stiffness was calculated using the oscillometric waveform envelope. The OMWE was plotted against *P*_*t*_. The signal was then numerically integrated, resulting in an S-shaped volume-transmural pressure (*V* -*P*_*t*_) plot. To fit the data into a descriptive function, we used nonlinear regression:

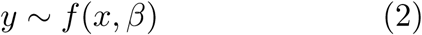

where *x* is an independent variable, *y* its associated dependent variable and *β* is a vector of parameters of a nonlinear function *f*. We fit the measured volume-pressure data to an inverse trigonometric function:

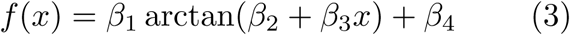

where *β*_1…4_ are the parameters computed using non-linear fitting and variable *x* is transmural pressure *P*_*t*_. Since *β*_2_ describes the slope of the curve, it is used as the arterial stiffness index (ASI).[16]

We used the oscillometric waveform to detect AF. This was done during BP computation after fitting the 8th order polynomial curve to the found peaks. The absolute difference, or error, between the found peak and the polynomial fit curve was calculated for each peak. The mean error was calculated for each measurement and compared with a threshold to decide between AF and sinus rhythm. The principle of this is shown in Figure 7.

### 2.3 Human studies

We recruited 32 volunteers (age: *μ* = 47 years [range: 24 to 83 years], 5 women) for measurements using the proposed device and manual auscultation. One subject was excluded due to severe nail clubbing. Six subjects having persistent atrial fibrillation were excluded from the BP validation data set, but were used to validate the detection of AF. The validation study was carried out at Turku University Hospital. Subjects were placed in a supine position to ensure that both for the finger device and the arm cuff were at the same level. Subjects were also asked to relax and wait 10 minutes before the first measurement to ensure that BP was stable. The finger probe was placed on the index finger and an arm cuff connected to a mercury column sphygmomanometer was placed around the upper arm. Manual auscultation was performed by two trained observers, and the maximum acceptable difference between the readings from the observers was 4 mmHg for both SBP and DBP. After an initial test measurement, three measurements were taken in a cyclical manner one device after another, both with the finger device and by auscultation. Data is displayed in Bland-Altman and correlation plots for SBP, MBP and DBP individually[1].

In order to broaden the BP range, we recruited 9 individuals (age: *μ* = 48 [range: 25 to 78 years], 4 females) to perform a hydrostatic challenge. Four of the subjects were on BP lowering medication. We measured BP using our device and a clinically validated wrist-worn BP cuff (Omron R2, Japan) [27]. The wrist cuff uses the oscillometric method for the acquisition of BP. Both instruments were worn in the same arm and held 10 cm below and above heart level resulting in a total of 18 data points of SBP, MAP and DBP. This results in a change of approximately 15 mmHg between the two levels. Three consecutive measurements were taken for both hydrostatic levels. Three of the subjects measured in this group were also in the auscultated group, but the measurements were taken over 6 months apart.

In addition, we took a series of subsequent measurements to study the device’s capability to take repeated measurements. The study was performed on one subject over approximately 20 minutes while comparing our device to a continuous reference. We used a volume clamp device CNAP 500 (CNSystems, Austria) as a reference [11]. CNAP 500 is capable of outputting continuous BP data including SBP, MAP and DBP as well as the full cardiac waveform. The reference device’s cuffs were placed on the middle and ring fingers ipsilaterally and our device was placed on the index finger. The reference device was calibrated using the built-in brachial sphygmomanometer at the beginning of the measurement and again at 15 minutes. A total of 9 measurements were taken with our device, approximately once every two minutes. A timestamp was set on the reference device after each oscillometric measurement taken on our device. As the oscillometric sweep takes approximately 20 s, the reference BP values were averaged over 20 s for each measurement. We computed ((*Mean* ± *SD*) mmHg) for the measurements.

PWV was recorded from 18 subjects using SphygmoCor XCEL (Atcor Medical, Australia) and XCEL 1.3 data acquisition software [7]. The patients remained in supine position while a femoral artery cuff was wrapped around the thigh. A pen-like tonometer was used to acquire the carotid waveform. The distance between the cuff and the carotid artery via jugular notch was measured and used to calculate the PWV. An average of three consecutive measurements was used for data analysis.

The measurements were conducted according to the Declaration of Helsinki guidelines with the permission of Ethical Committee of the Hospital District of Southwest Finland and National Supervisory Authority for Welfare and Health[2].

## 3 Results

### 3.1 Blood pressure

Accuracy of the blood pressure measurement was verified by combining the two data sets collected: the auscultatory set (n=25) and the hydrostatic challenge set (n=18). A total of 43 pairs of BP values were analyzed. The blood pressures measured ranged from 96 to 152 mmHg and from 50 to 96 mmHg for SBP and DBP respectively. We calculated ((*mean* ± *SD*) mmHg) for SBP, MBP and DBP, resulting in ((−4.9±9.5) mmHg), ((−1.2 ± 3.9) mmHg) and ((−4.0 ± 4.4) mmHg) respectively. The patient-specific method for computing SBP performed better ((−3.5 ± 8.4) mmHg) than the standard method. Mean and diastolic pressure values comply with the international standards for non-invasive BP monitors. Standard deviation for SBP (8.4 mmHg) was 0.4 mmHg higher than the AAMI standard threshold (8 mmHg) [30]. However, the method for computing SBP was very simple and we believe that using more sophisticated computation techniques, the standard could be met even for SBP. Moreover, the accuracy of MBP is suitable for longitudinal bedside BP monitoring.

### 3.2 Repeatability

We wanted to evaluate the device’s capability for long term monitoring. We took 9 consecutive measurements from the same subject over a 20 min period. Consecutive measurements taken with our device were very close to the reference device and followed the changes in BP correctly. Comparison of the repeated measurements to the reference device resulted in SBP of (−6.9 ± 4.7) mmHg, MBP of (0.6 ± 4.0) mmHg, and DBP of (1.6 ± 3.4) mmHg. The entire measurement is shown in Fig. 5.

**Fig. 1.**
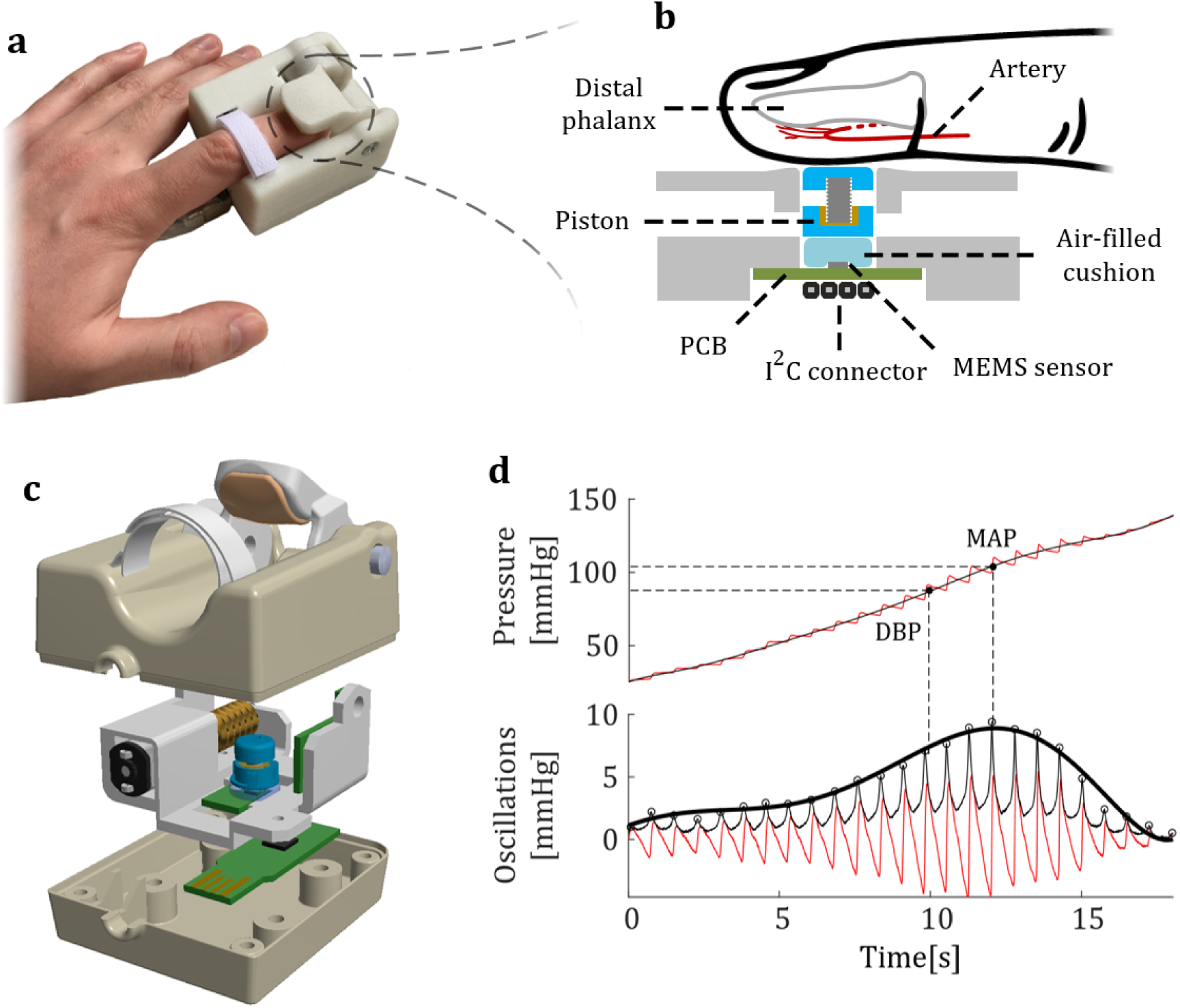
Instrument overview. a) Photograph showing the device in use. b) Cross-sectional view of the sensor setup. c) 3D model of the device. d) Oscillometric method. The pressure ramp signal is filtered with a pass-band filter (1 - 10 Hz) and Hilbert transformed. The peaks, or oscillations, are detected, and a polynomial curve is fitted to the peaks. The resulting bell-shaped curve is called an oscillometric waveform envelope (OMWE). Mean arterial pressure (MAP) is found at the point on the pressure curve that corresponds with the maximum of OMWE. DBP is found on the left side of the OMWE at 80% of maximum pressure. SBP is computed from DBP and MAP.

**Fig. 2.**
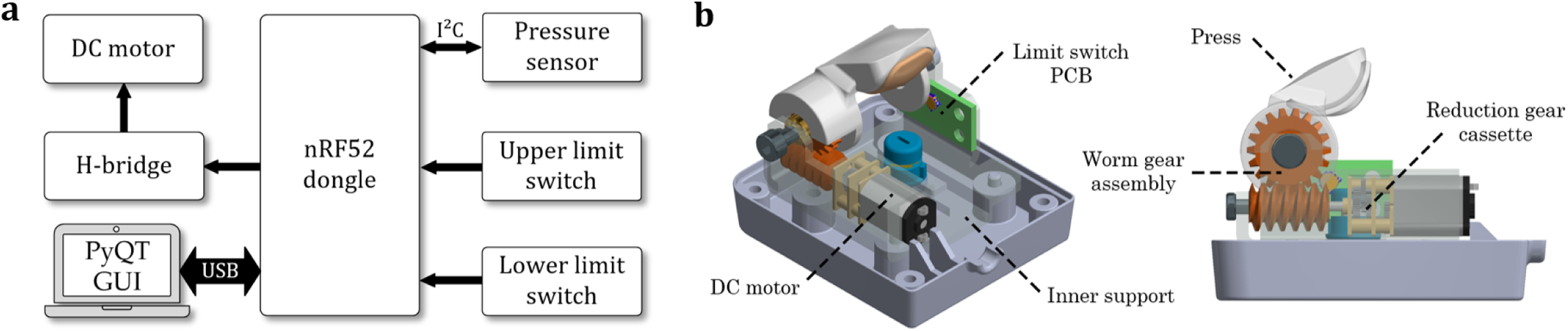
Description of the system. a) Block diagram of the system. An ARM Cortex M4 based MCU is used as the controller unit. The MCU is connected to a laptop running a Python GUI for data logging. b) Inner mechanics illustrated in a 3D view. A DC motor is connected to a worm gear assembly through a reduction gear cassette. This rotates a hinge-type press that is used to apply pressure to the finger. A pair of micro limit switches assembled on a custom circuit board is used to limit the motion.

**Fig. 3.**
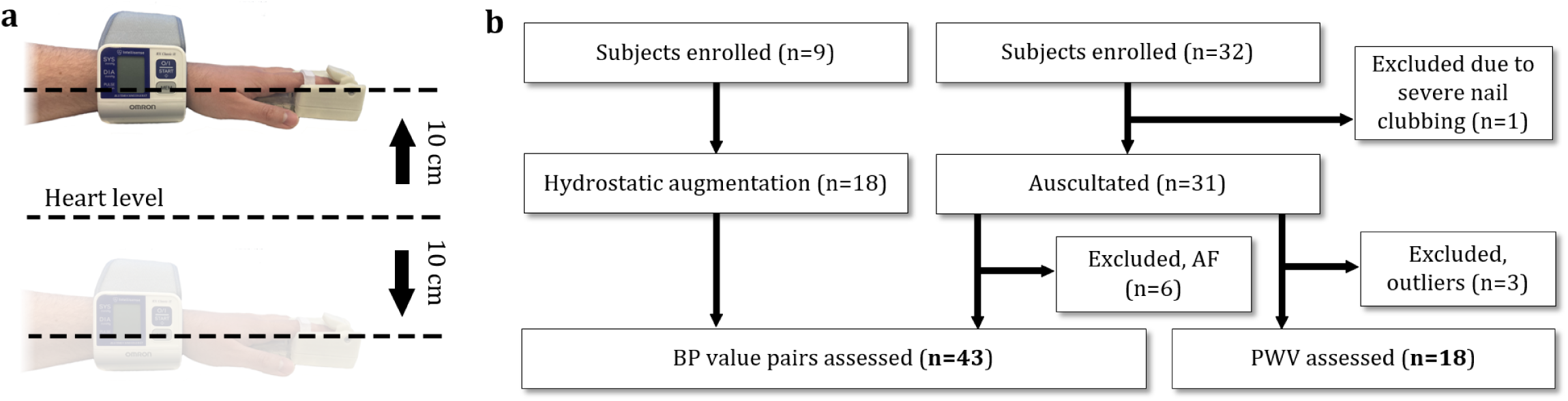
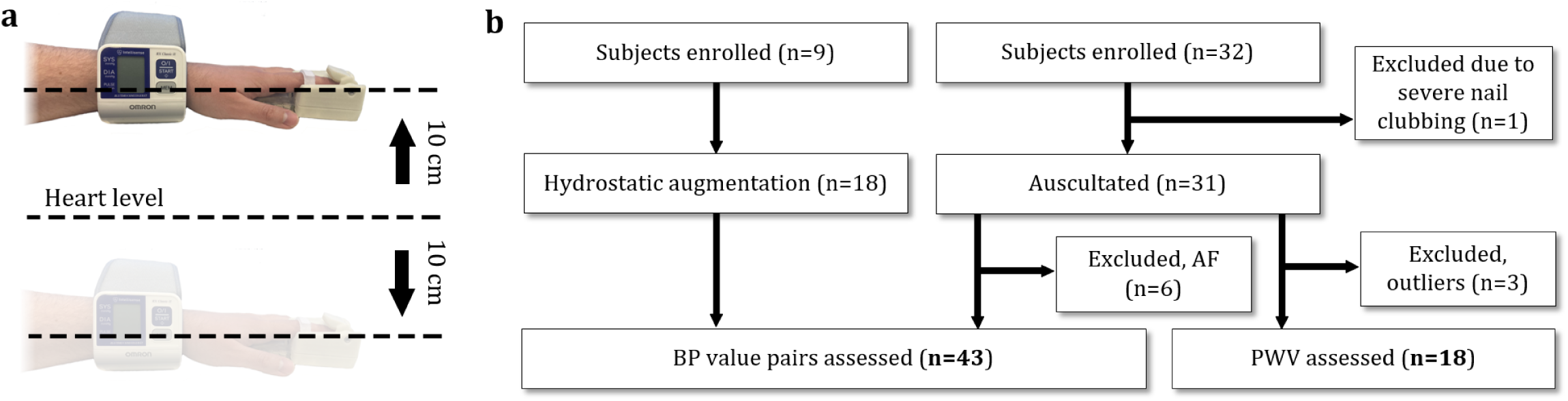
a) Hydrostatic challenge. The subjects wore the finger device and a reference wrist BP monitor on the same hand. Three measurements were taken with the hand raised above and lowered below heart level, causing a change in BP. b) Clinical trial flow chart.

**Fig. 4.**
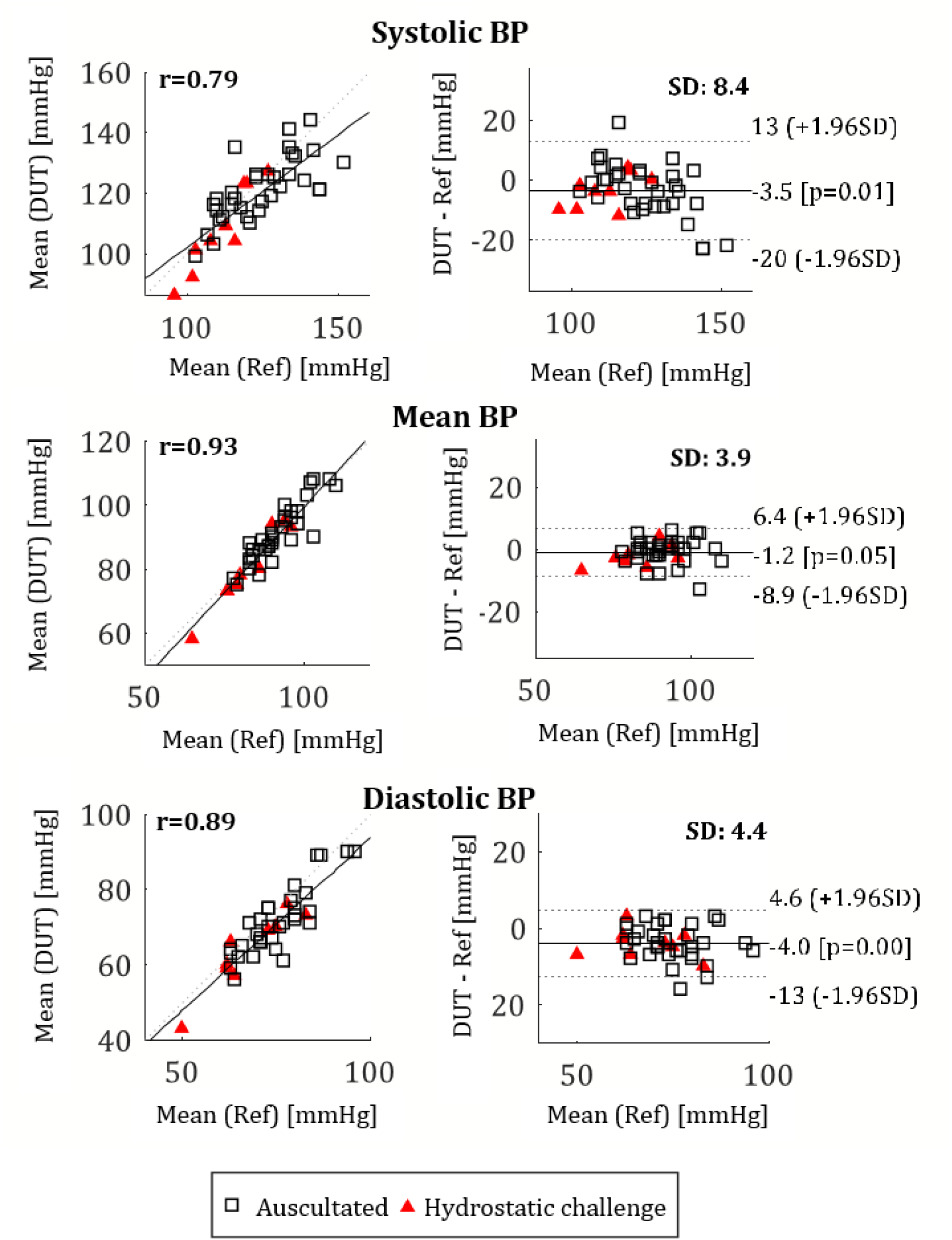
Bland-Altman and correlation plots for SBP, MBP and DBP. The two data sets, auscultated and hydrostatic challenge, are plotted with different markers.

**Fig. 5.**
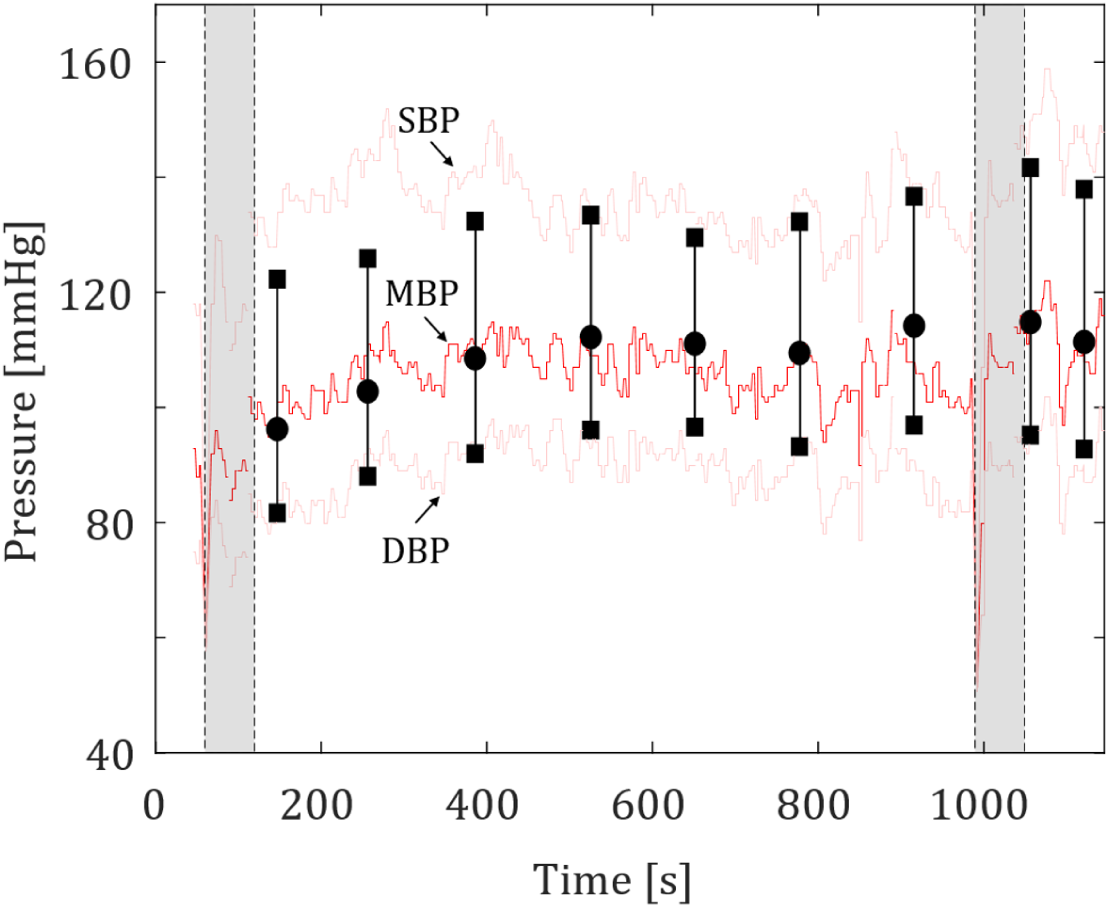
Repeatability study. Oscillometric measurements (shown as black vertical lines and markers) were taken with the finger device approx. every 2 minutes. The red and light red lines represent the continuous BP taken with the reference device CNAP 500.

**Fig. 6.**
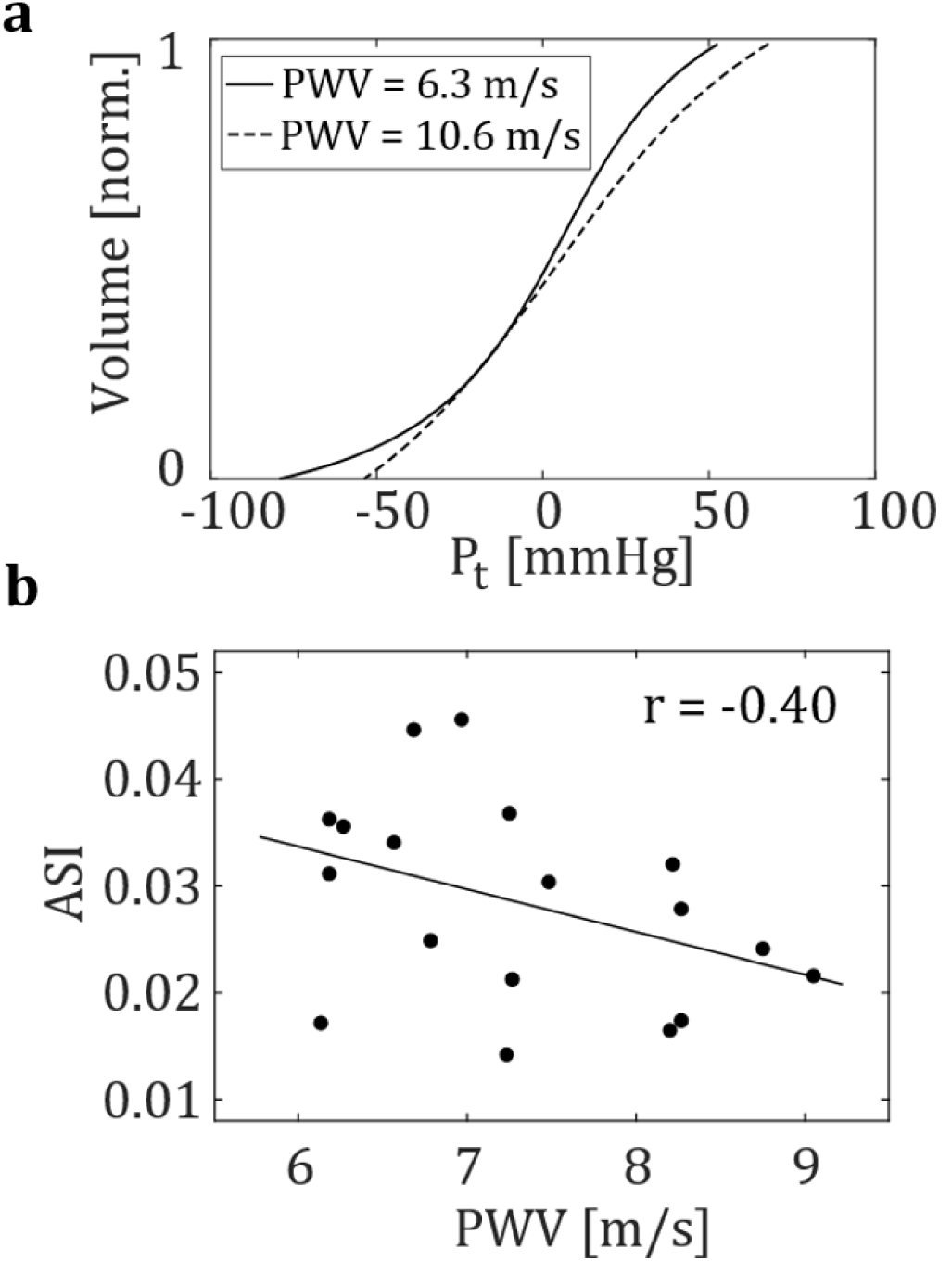
a) Example *V* -*P*_*t*_ plots estimating arterial compliance for a young subject with low carotidto-femoral PWV (6.3 m/s) and an elderly subject with high PWV (10.6 m/s). The slope of the curve in the young subject’s case is higher, resulting in higher ASI. b) Figure showing the negative correlation between ASI and PWV.

### 3.3 Arterial stiffness

Arterial stiffness was evaluated by analyzing the shape of the oscillogram acquired by the finger device, resulting in an arterial stiffness index (ASI). The ASI describes the compressibility of the artery when external pressure is applied to it. This is characterized by the slope of the curve in the *V* -*P*_*t*_ plot. The computed ASI was compared with various other parameters. PWV and ASI showed a negative correlation (r=-0.4), which is comparable to the results presented in the literature[16]. This suggests that individuals with high PWV readings have lower ASI, thus less compliant arteries. Individuals with lower PWV seem to have more variation in ASI with high and low ASI values. Similar phenomenon was noticed when we compared ASI and age. Younger subjects (*<*40 y) tend to have higher variation in ASI than older ones (*>*60 y). However, the younger group shows a 40% higher mean value than the older group. In interpreting the results between PWV and ASI, it is important to understand that PWV is dependent on two intrinsic variables: arterial stiffness and BP. Due to this, PWV and ASI do not have a linear relationship [10]. As can be concluded from the results shown in Subsection 3.1, ASI does indeed hold BP information and can be used as an additional parameter for computing BP.

### 3.4 Atrial fibrillation detection

We wanted to both demonstrate the device’s capability of detecting AF and exclude these patients from the validation data set, since AF is known to affect oscillometric BP measurements significantly. AF was found on six subjects. The mean error from OMWE in mmHg was computed for each subject. In AF, the pulse pressure varies randomly, resulting in deformation of the oscillometric envelope. Single pulses can be higher or lower than the OMWE, increasing the mean error. A 0.8 mmHg threshold was used to differentiate AF from sinus rhythm. Figure 7 shows that AF cases can be classified using a simple threshold.

**Fig. 7.**
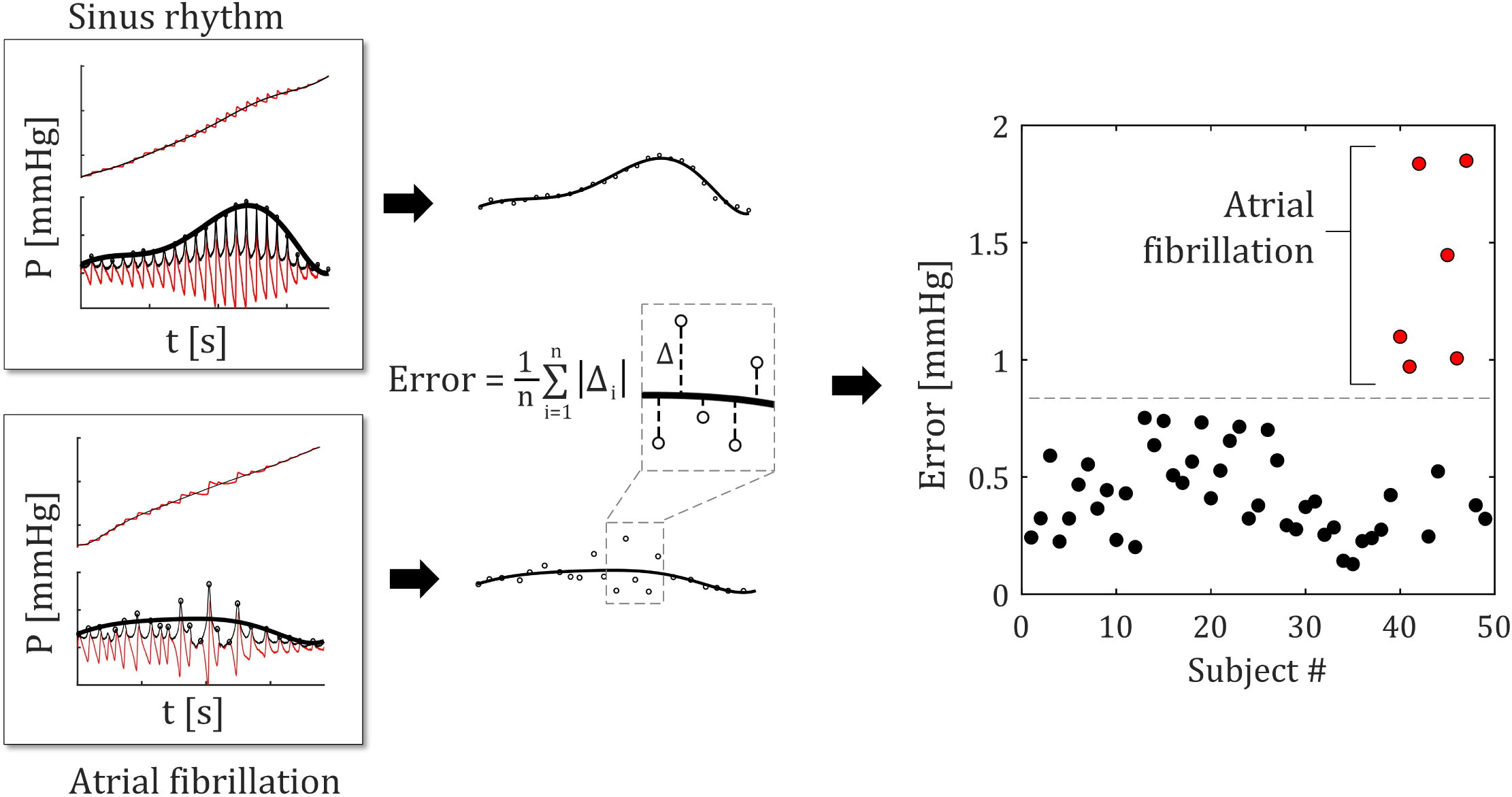
Atrial fibrillation detection. The difference between the oscillometric envelope and the amplitude of each peak is used to calculate the mean error. The algorithm was able to differentiate AF from sinus rhythm using the error between oscillometric envelope and pulse peaks.

## 4 Discussion

We presented a miniaturized finger probe for measuring BP, assessing arterial stiffness and detecting atrial fibrillation. In this study, the sensor technology introduced in our previous article was integrated into a pulse-oximeter-style wearable probe [23]. This allows the device to be discrete and unobtrusive while making integration to existing patient monitors possible. We validated the the device in a clinical trial, comparing the proposed device with auscultation the gold standard method for non-invasive BP measurement. In an attempt to improve the credibility of the validation, we introduced additional high- and low-BP values into the data set via hydrostatic challenge. This way we were able to record more high and low pressures without having to recruit subjects having such BP values. The results for MBP and DBP fulfill the bias and precision criteria set by AAMI, but the standard deviation for the SBP are slightly above the standard threshold (0.4 mmHg above the limit). The accuracy could be improved by more sophisticated algorithms and better positioning of the finger. One possibility is either to use a personalized molded insert or an adjustable back piece to make sure the device geometry is optimal for each individual. This would be viable especially for long-term use. The study differs from standard protocol in population size and BP range. Our study size (n = 43) satisfies the criteria for the number of subjects set by ESH (n=33) but not by AAMI (n=85). In addition, we used the hydrostatic challenge to increase the data set, which is not a method recommended in any of the standards. Not all of the study population requirements were met either. The subjects recruited in the study were either normotensive or had their hypertension under control. We were able to broaden the BP range with hydrostatic challenge, but we still miss the very high BP of *>*160 mmHg for SBP and *>*100 mmHg for DBP. Arterial stiffness was evaluated by analyzing the shape of the oscillogram. Our measurements suggest that ASI can be used as an additional parameter for computing BP. We found a negative correlation between ASI and carotid-to-femoral PWV, similar to the findings in the literature[16].

Age and ASI were also found to correlate. ASI could possibly be used as a feature in machine learning applications for more pervasive hemodynamic monitoring. We were also able to distinguish AF cases from sinus rhythm cases using simple metrics derived from the oscillometric envelope.

The finger-pressing system has a clear advantage over cuff-based systems: One device is suitable for all finger sizes. Brachial and finger cuff devices often have different cuffs for small, medium, and large anatomies. The study subjects also noted that the finger device produced much less discomfort than a traditional brachial cuff. Especially in the case of hypertensive patients, the pressure required for complete brachial occlusion can cause significant pain. Compression applied on the fingertip causes less distal tissue occlusion compared to brachial, radial, and proximal phalanx cuff occlusion. Furthermore, taking short oscillometric measurements periodically is much less uncomfortable than having to maintain partial occlusion continuously, which is the case in volume-clamp methods [12]. Another significant advantage to the volume-clamp method and most wearable devices is that our approach does not need any form of calibration, which is usually done by brachial cuff oscillometry.

There are also limitations to the technology. In several cases, the subject had cold fingers, which significantly affected the quality of the signal. One subject had undergone surgery on his left arm, making it difficult for him to hold his hand in the correct position. Additionally, for several subjects, the subject’s finger had to be repositioned several times to ensure good signal quality and coupling of arterial pulsation to the sensor. This suggests that the sensor is quite sensitive to misplacement. However, when correctly positioned, the measurements had high repeatability, as demonstrated in Subsection 3.2, which is crucial for long-term bedside monitoring.

Additionally, future directions could include a more exhaustive study on pseudo-continuous BP monitoring. An ideal setup would include an invasive reference e.g. during a surgical operation. In our previous publication, we demonstrated that using fingertip tonometry, we were able to acquire high resolution BP waveform similar to invasive BP [13]. The combination of periodic oscillometric measurements and pulse waveform could be of clinical use. Another interesting topic is to study how the device performs in nocturnal BP monitoring, possibly including optical oxygen saturation sensing. Nocturnal hypoxemia, that is, a decrease in peripheral oxygen saturation during sleep is a marker of adverse conditions, such as chronic obstructive pulmonary disease [5] and obstructive sleep apnea. Furthermore, it has been reported to inversely correlate with morning surge of BP, which, on the other hand, is an important diagnostic tool in the management of hypertension [14, 15]. Making the device wireless and battery-operated would enable convenient data collection in nursing homes or even in a home environment. The current enclosure still has room for a small lithium-polymer battery as well as a charging circuit, and the microcontroller used has Bluetooth radio communication capabilities. However, this is more of a novel engineering challenge. Compared to a bulky BP device, a small portable device would be easier for the healthcare professional to carry on the hospital ward.

## Data Availability

All data produced in the present study are available upon reasonable request to the authors

## Author Contributions

Conception and design: T.P., T.N., I.K. and M.K., System development: T.P. and J.S., Collection and assembly of data: T.P., J.S., T.N., I.K. and M.K., Analysis and interpretation of the data: T.P., Drafting of the article: T.P., J.S. and M.K., Obtaining funding: M.P., T.K. and M.K.

